# Knowledge, attitude and willingness to accept the RTS,S malaria vaccine among mothers in Abuja Nigeria

**DOI:** 10.1101/2020.12.03.20242784

**Authors:** Tolulope O. Musa-Booth, Blessing E. Enobun, Adewumi J. Agbomola, Clive J. Shiff

**Affiliations:** Department of Molecular Microbiology and Immunology, Johns Hopkins Bloomberg School of Public Health, 615 N. Wolfe St, Baltimore, MD, 21205 USA; University of Maryland, Baltimore, USA; National Malaria Elimination Programme, Abuja, Nigeria

**Keywords:** knowledge, attitude, malaria prevention, RTS,S malaria vaccine, vaccine acceptance

## Abstract

**Objective:** To assess mothers’ knowledge, attitude, and practices to malaria and its prevention as well as mothers’ willingness to accept the RTS,S/AS01 malaria vaccine.

**Methods:** A cross-sectional study design was used to administer questionnaires to 180 mothers within six public secondary health facilities in Abuja, Nigeria. Regression analyses were used to assess the effect of maternal demographics and exposure to malaria related messages on the outcome measures.

**Results:** Knowledge of malaria preventive measures was approximately 59% (124/180). Multivariate logistic regression analyses indicated ethnicity, education and number of messages or training on malaria were associated with mother’s knowledge of 3 or more preventive measures against malaria. About 30% (36/180) of respondents were aware of malaria vaccines but only four percent (7/180) had heard of RTS,S. Young maternal age (OR, 2.4; p = 0.03), self-employment (OR, 2.55; p = 0.04) and formal employment (OR, 3.74; p = 0.03) were associated with no prior knowledge of malaria vaccine. Ninety-one percent (163/180) had positive attitude to malaria vaccine and 98% (176/180) were willing to allow their child(ren) to be immunized with RTS,S despite the efficacy of the vaccine, route of administration and number of doses.

**Conclusion:** Knowledge of malaria preventive measures does not correlate with knowledge of RTS,S. Although willingness to accept RTS,S is high, consistent targeted messaging on RTS,S would be required to improve community knowledge and attitude prior to implementation to ensure maximum uptake.

## INTRODUCTION

WHO burden estimates that 92% malaria cases and 93% deaths from malaria in 2017 were seen in Africa.^1^ Countries in sub-Saharan Africa and India account for 80% of the global burden and deaths.^1^ From 2015-2017, there was no significant reduction in the malaria cases recorded globally. Although, the 2020 world malaria report shows that there was a reduction in number of deaths in the high burden to high impact countries, there was still an increase in number of cases.^2^ Nigeria, which is malaria endemic and accounts for 23% of the global burden, also reported an increase in malaria cases in 2019^2^ despite the investments and interventions in place to control the disease. Nigeria also accounted for 19% of all global malaria deaths.^1^ Based on the last National Malaria Indicator Survey (NMIS) done in Nigeria, *Plasmodium falciparum* malaria accounts for majority of outpatient visits (60%) and admissions (30%).^3^ It contributes about 11% of maternal mortality, 25% of infant mortality, and 30% of under-5 mortality^3^. Approximately 110 million clinically diagnosed cases of malaria and nearly 300,000 malaria-related childhood deaths occur each year.^3^ Malaria remains a major public health problem in Nigeria, weakening the health system and causing losses of about 40% annually in the economy’s gross domestic product (GDP) to the tune of 480 billion naira in out-of-pocket treatments, prevention costs, and loss of man hours.^3^ Nearly three quarters of the 3.1 billion U.S. dollars invested in malaria control and elimination globally in 2017 was spent in Africa.^1^

To meet the WHO global technical strategy targets^4^ and the sustainable development goal 3.3,^5^ more funding will be required for research, and development of malaria vaccines in addition to the interventions currently in place. With the COVID-19 pandemic taking a toll on health systems worldwide, it will be a challenge to achieve the set targets for malaria control and elimination but with rising number of cases and deaths, vaccine science has been brought to the fore. Over 30 *P. falciparum* malaria vaccine candidates are either at advanced preclinical or clinical stages of development but only RTS,S/AS01 vaccine, has passed phase three clinical trial.^6^ Results showed that the vaccine was effective especially when used in conjunction with other preventive measures.^7, 8^ A pilot implementation study was subsequently recommended by WHO and is ongoing in Ghana, Malawi and Kenya to evaluate how four doses of RTS,S can be implemented among 5-17month old children at first dose and the risk/benefits of the vaccine.^7^

There is very little data on the knowledge, attitude and practices of Nigerians or any society, to the potential RTS,S malaria vaccine. A study done in Ibadan, southwest Nigeria revealed that only 20% of caregivers had heard of malaria vaccine.^9^ Although approximately 87% were willing to accept the vaccine, almost 50% of the caregivers did not think the vaccine would be accepted if it was not given orally.^9^ Another study done in Owerri west, southeast Nigeria showed approximately 48% knowledge of malaria vaccine, 88.2% positive perception of the vaccine and 95.6% willingness to accept the vaccine.^10^ Our structured study represents a cross section of maternal population in the city of Abuja, northcentral Nigeria to assess their understanding of the role of malaria vaccines.

## MATERIALS AND METHODS

### Study Design and Population

Following the Johns Hopkins institutional review board and FCT Health Research Ethics Committee approval, a cross sectional study design was adopted to assess knowledge, attitude, and practices to malaria, malaria prevention and RTS,S vaccine among 180 consenting mothers in six public secondary health facilities within Abuja (Figure 1). Mothers’ willingness to allow their children to receive RTS,S vaccine was also assessed as well as mothers’ socio-demographic characteristics using a validated semi-structured interviewer-administered questionnaire. The study was conducted between February and March 2020.

**Fig.1:**
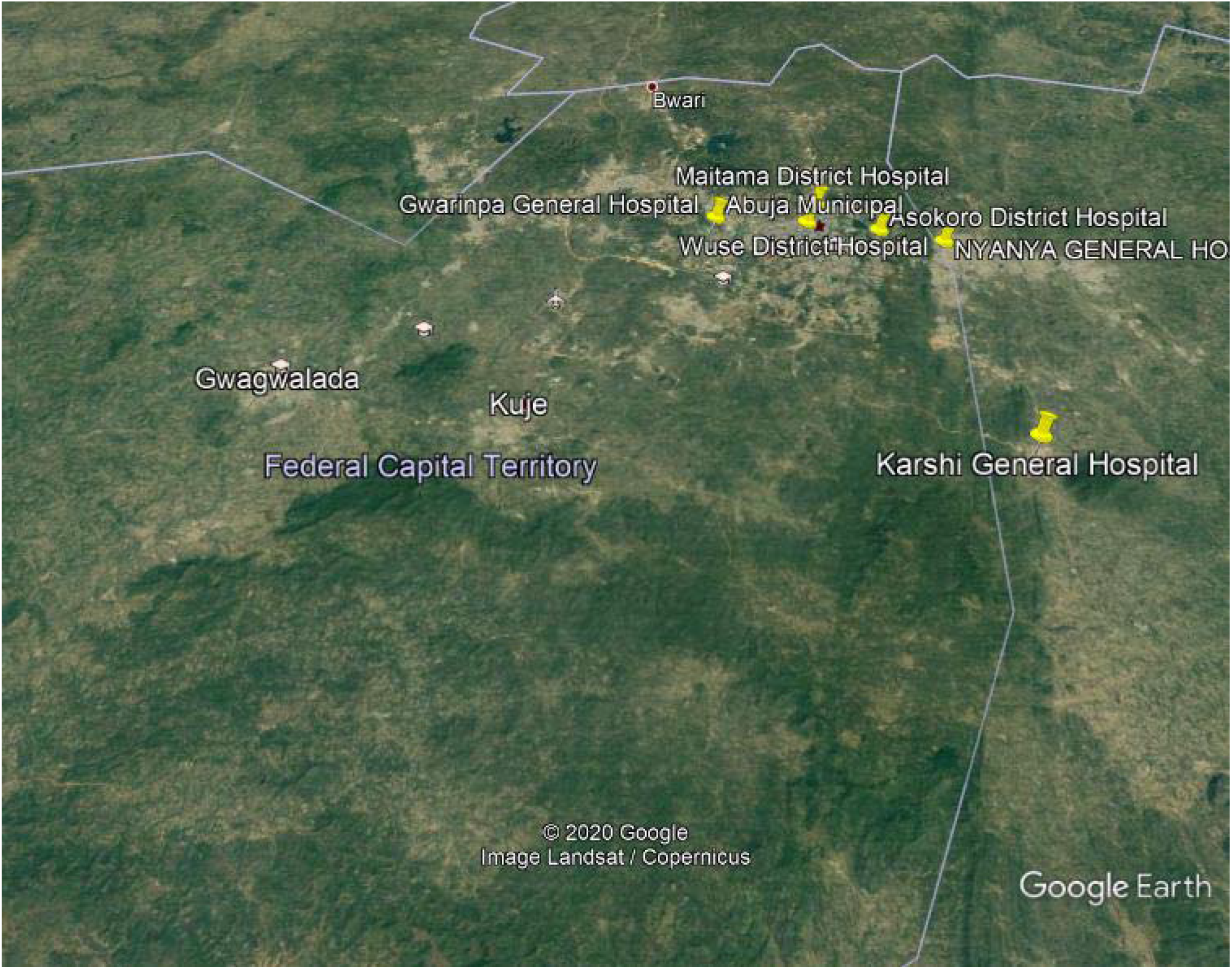
Geospatial Map of Public Secondary Health Facilities in AMAC.

### Statistical Analyses

Data was analyzed using Stata/MP 15.0 (StataCorp. 2017. *Stata Statistical Software: Release 15*. College Station, TX: StataCorp LLC). Descriptive statistics was used to summarize the socio-demographic characteristics. Mean and standard deviation were employed to summarize continuous data. Counts and percentages were utilized to summarize categorical variables. Potential predictors (independent variables) were assessed for a statistical association with the outcome variables (the dependent variables) using univariate analysis and multivariable logarithmic regression analysis with inclusion of 0.10.

### Outcome Variables

Knowledge of malaria preventives measures was coded as a dichotomous variable: less than three preventive measures vs three or more preventive measures, to assess mothers’ knowledge on malaria prevention. Knowledge of malaria vaccine was measured as a dichotomous variable: yes or no. Mothers’ attitude to malaria, its prevention and malaria vaccine was assessed using a malaria attitude score. The malaria attitude score was a sum of the Likert scale values of the six questions on attitude in section D of the questionnaire. The scoring on the Likert scale for three questions (avoid contact with someone who has malaria, not immunized with malaria vaccine and immunization with malaria vaccine) was reversed to tally positively with the other three questions. The minimum malaria attitude score a respondent could attain was six points and the maximum was 30 points. A cut-off of 24 points was selected as the threshold which corresponds to a score of four on each of the six questions on attitude. A score of 24 points and above represented good attitude and a score below 24 points, poor attitude on malaria and malaria prevention. Mothers’ practices regarding malaria vaccination was measured by willingness to allow child to be immunized with malaria vaccine, bring child for vaccination, immunize child by injection and use of other preventive measures after vaccination. All four outcomes were dichotomized as yes, no responses.

## RESULTS

### Socio-demographic Characteristics

The average age of respondents was 30 years with a vast majority of mothers being married (96%). 58% of respondents had post-secondary education and a little over half of the respondents were self-employed (54%). Mothers were predominantly Christian (86%) and the three main ethnic groups were well represented although Igbos made up majority of respondents (24%). Some of the other tribes captured include Idoma, Igala and Edo.

### Knowledge of malaria, its prevention and RTS,S vaccine

All (180/180) respondents had heard of malaria. 81% (145/180) of respondents knew mosquito bite causes malaria, 59% (124/180) knew one or two malaria preventive measures and 94% (170/180) knew everyone was at risk of infection. 28 of 180 (16%) respondents had not seen any message nor received any training on malaria, 46% (82/180) saw messages or received training on malaria from one or two sources, 31% (56/180), three to five sources and seven percent (12/180), more than five sources. Majority of respondents (92%; 166/180) knew malaria could be fatal if untreated, however, only 30% (36/180) of respondents were aware of malaria vaccine. Five percent (8/180) knew what a malaria vaccine was used for and only four percent (7/180) had heard of RTS,S.

Multivariable logistic regression analyses indicated ethnicity, education and number of messages or trainings on malaria were associated with mother’s knowledge of three or more preventive measures against malaria. Igbos and other ethnic groups were 70% less likely (OR, 0.27; 95% CI, 0.08-0.87; p=0.03; and OR, 0.27; 95% CI, 0.10-0.77; p=0.01 respectively) to know three or more preventive measures against malaria compared with Hausas. Yorubas like Hausas had knowledge of at least three preventive measures against malaria (OR, 0.47; 95% CI, 0.12-1.76; p=0.26). Post-secondary respondents compared with pre-secondary respondents were 87% less likely (OR, 0.13; 95% CI, 0.02-0.90; p=0.04) to know at least three malaria preventive measures. There was marginal significance in the knowledge of at least three malaria preventive measures between respondents who have attained secondary (OR, 0.16; 95% CI, 0.02-1.11, p=0.07) or other educational status (OR, 0.04; 95% CI, 0.001-1.13; p=0.06) compared with those with pre-secondary education. Respondents who had seen or received at least three messages and trainings on malaria were 8.2 times more likely to be aware of at least three malaria preventive measures (95% CI, 3.77-18.0, p < 0.001) (Table 2).

**Table 1:** Socio-demographic characteristics of respondents Characteristics Respondents (N=180)

| **Characteristics** | **Respondents (N=180)** |
| --- | --- |
| **Age (years)** | n = 179 |
| **Min-Max** | 16 – 49 |
| **Mean (SD)** | 30 (5.6) |
| **Marital Status n (%)** |  |
| **Single** | 7 (4) |
| **Married** | 173 (96) |
| **Religion n (%)** |  |
| **Christianity** | 154 (86) |
| **Islam** | 26 (14) |
| **Traditional** | 0 (0) |
| **Ethnicity n (%)** |  |
| **Hausa** | 30 (17) |
| **Igbo** | 44 (24) |
| **Yoruba** | 22 (12) |
| **Other** | 84 (47) |
| **Education\* n (%)** |  |
| **Pre-secondary** | 6 (3) |
| **Secondary** | 67 (37) |
| **Post-secondary** | 103 (58) |
| **Other** | 3 (2) |
| **Employment status** | n = 179 |
| **Not employed** | 37 (21) |
| **Self-employed** | 96 (54) |
| **Formally employed** | 42 (23) |
| **Other** | 4 (2) |

**Table 2:**
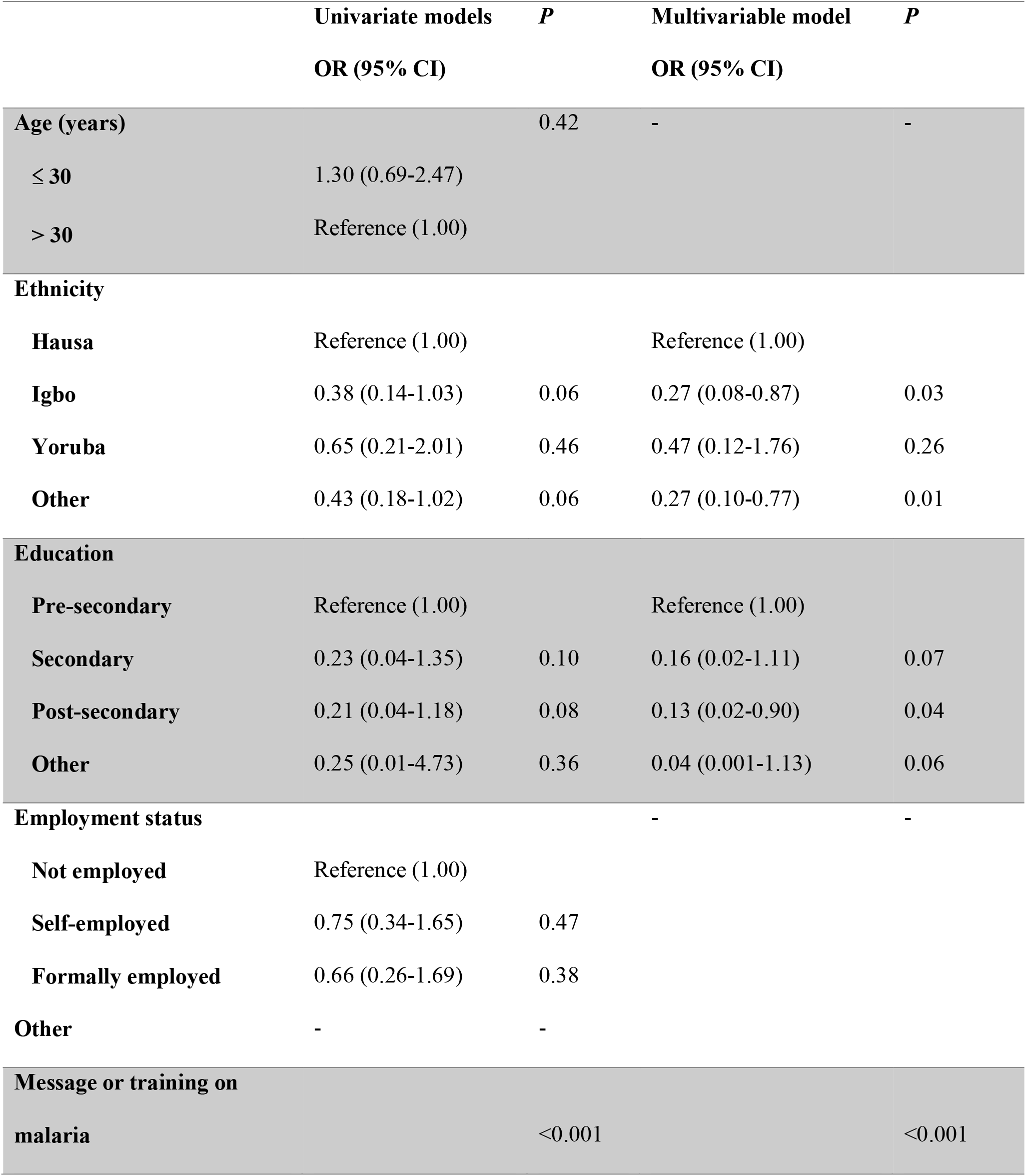
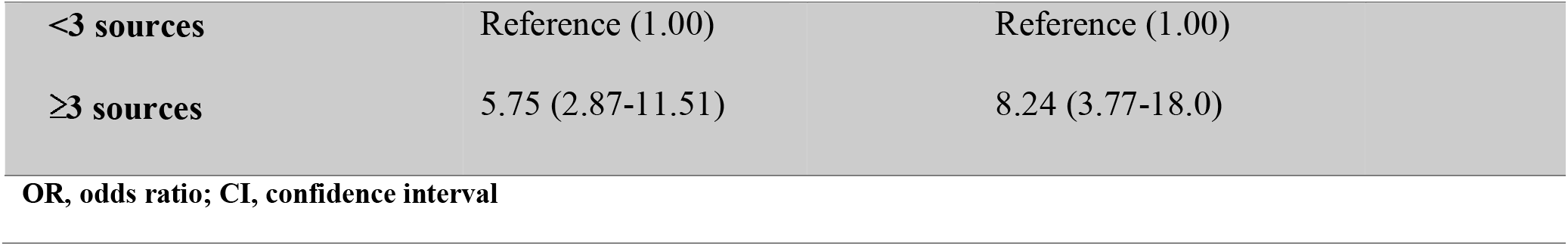
Factors associated with Mother’s Knowledge of three or more Malaria Preventive Measures.

Respondents 30 years and younger were more than two times more likely to be unaware of the existence of a malaria vaccine compared with older respondents (OR, 2.40; 95% CI, 1.09-5.28; p=0.03). Compared with unemployed respondents, self-employed (OR, 2.55; 95% CI 1.04-6.28; p=0.04) and formally employed respondents (OR, 3.74; 95% CI 1.17-11.99, p=0.03) were more than two times more likely to have no prior knowledge of malaria vaccine. No prior knowledge of malaria vaccine was similar in both the unemployed respondents and respondents with other forms of employment (OR, 1.94; 95% CI, 0.17-22.59, p=0.6). No statistically significant difference in prior knowledge of malaria vaccine existed between respondents with less than three sources of messages or training on malaria and those with at least three (OR, 1.97; 95% CI, 0.84-4.61; p=0.12) (Table 3).

**Table 3:**
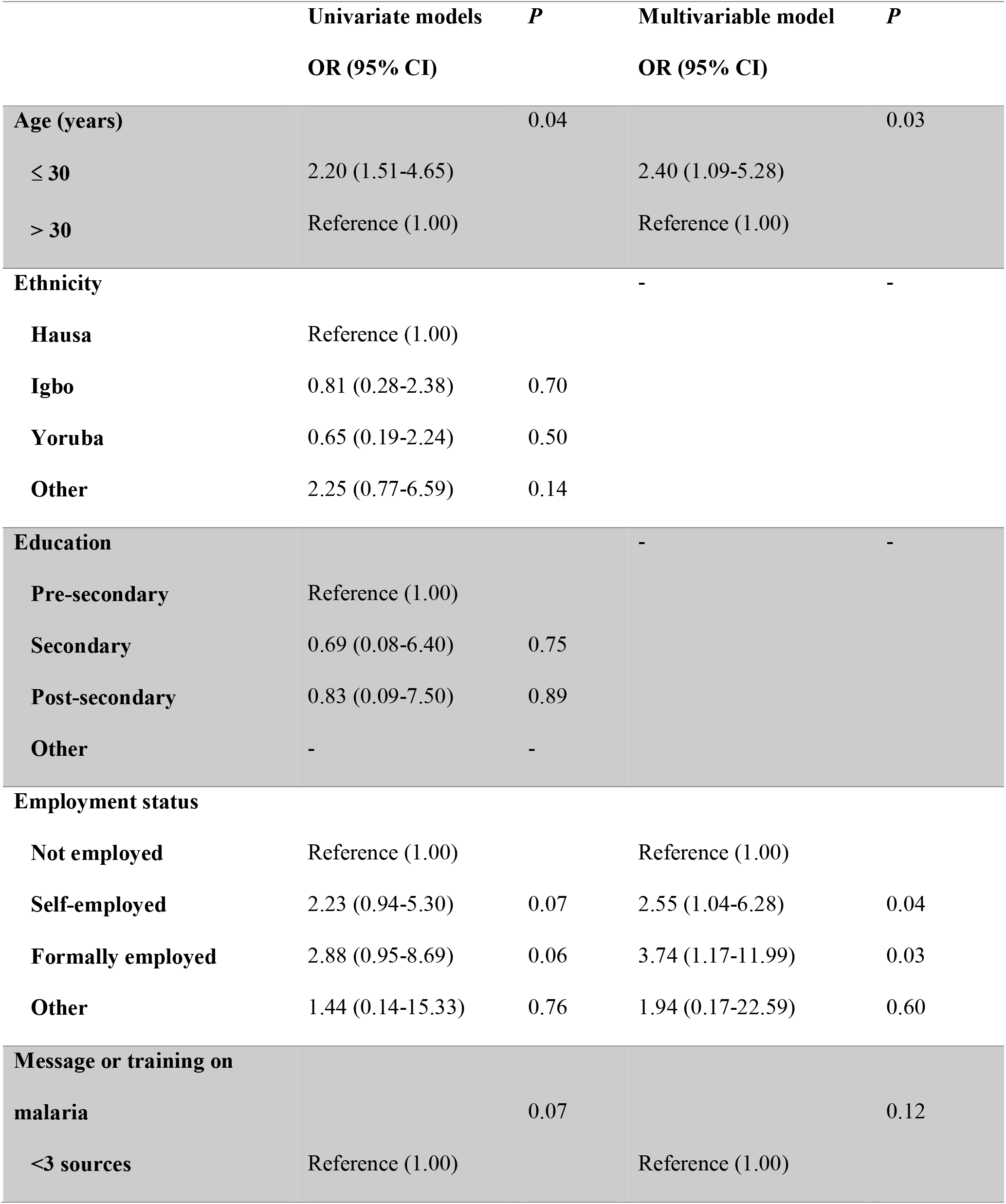
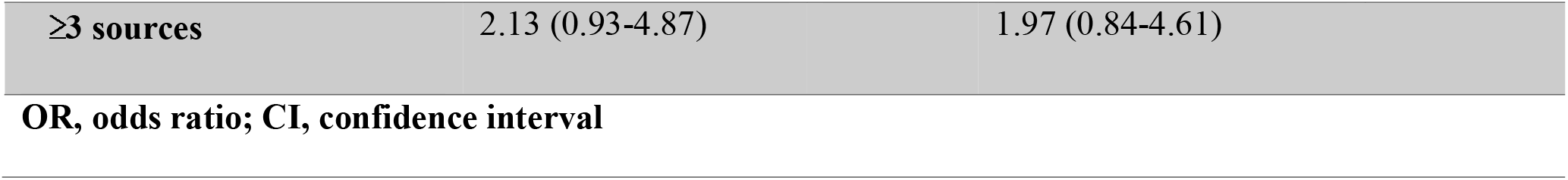
Factors associated with No Prior Knowledge of Malaria Vaccine.

### Attitude to malaria, its prevention and RTS,S vaccine

Majority of mothers (89%; 160 of 180) agreed or strongly agreed that malaria was a serious and life-threatening disease. 91% (163 of 180) of respondents agreed or strongly agreed that sleeping inside a mosquito net was one way to prevent malaria, 91% (163 of 180) also agreed or strongly agreed that vaccines are important in managing malaria and 21% (38 of 180) disagreed or strongly disagreed that immunization with malaria vaccine provided lifelong protection from malaria. 159 respondents (88%) disagreed or strongly disagreed that close contact with someone who had malaria should be avoided and 81% (146 of 180) also disagreed or strongly disagreed that anyone not immunized with malaria vaccine should be avoided.

There was zero percent floor and ceiling effects on the malaria attitude scale. We performed multivariable logistic regression analyses which revealed Hausas were more than three times more likely to have a low malaria attitude score compared with Igbos (OR, 3.14; 95% CI, 1.06-9.33; p=0.04). There were no statistically significant differences in malaria attitude score among Yorubas (OR, 1.85; 95% CI, 0.62-5.56; p=0.27) and other ethnic groups (OR, 1.19; 95% CI, 0.56-2.54; p=0.65) compared with Igbos. Respondents with less than three sources of messages or trainings on malaria were almost two times more likely (OR, 1.97; 95% CI, 1.04-3.73; p=0.04) to have a low malaria attitude score compared with those having three or more sources (Table 4).

**Table 4:**
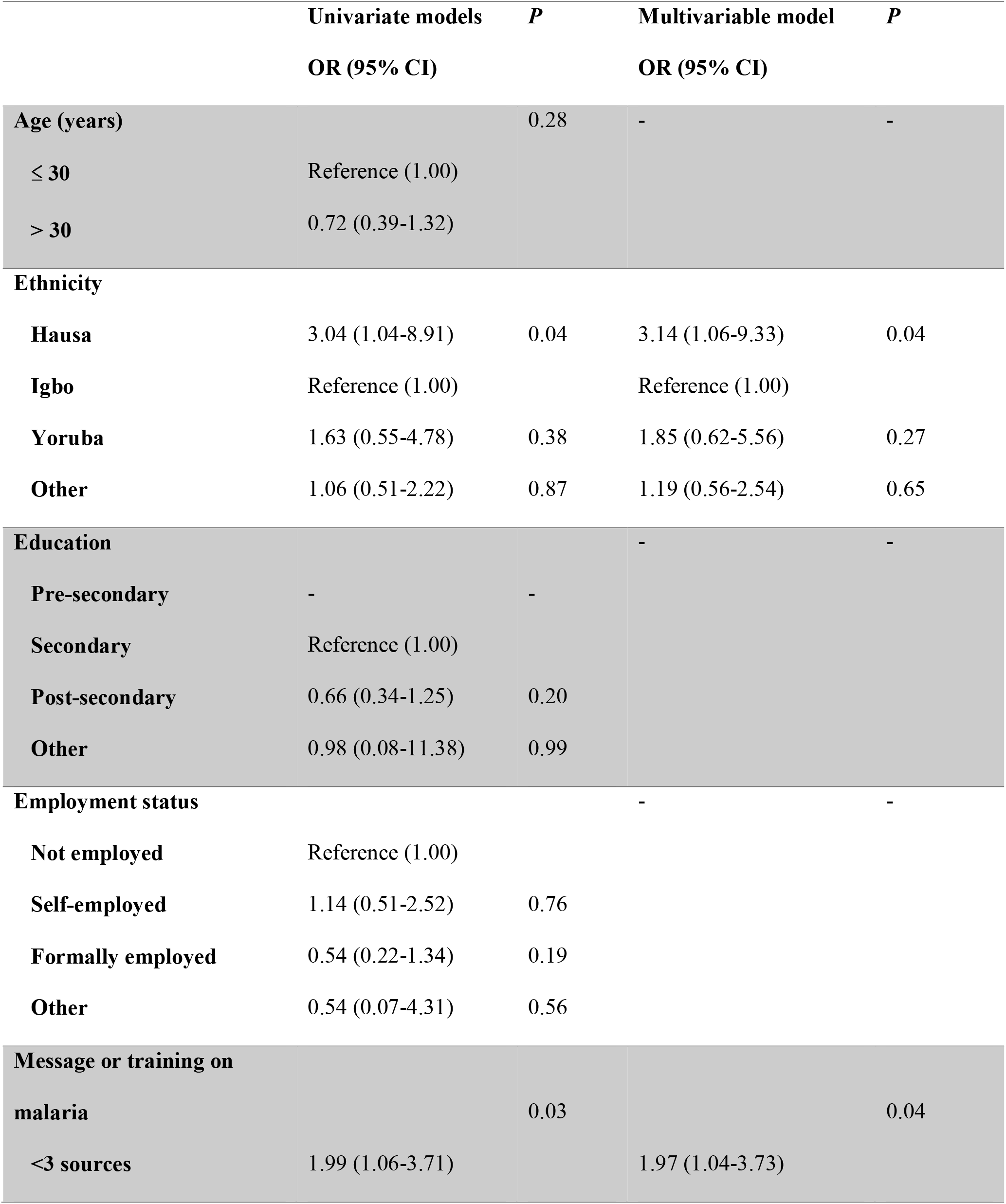
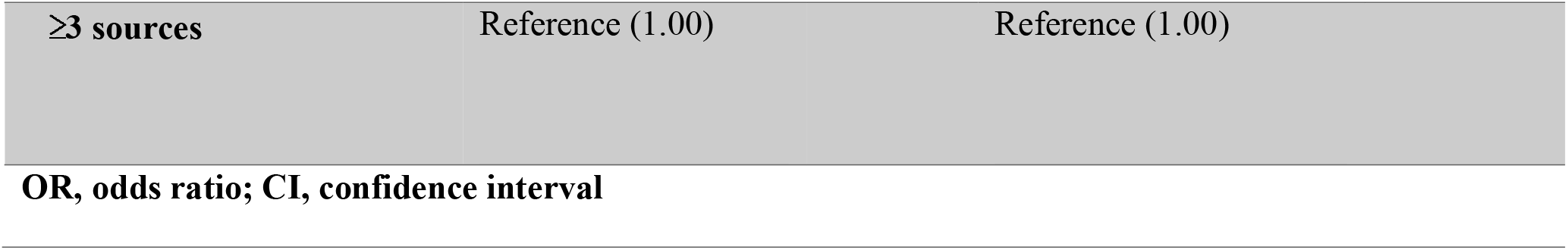
Factors Associated with Low Malaria Attitude Score (< 24 points)

### Practices regarding malaria prevention and vaccination

Most of the respondents (66%, 119 of 180) sometimes or always slept in insecticide treated nets and 80% (144 of 180) sometimes or always used anti-mosquito spray. 62% (112 of 180) of respondents sometimes or always cut bushes and grasses around the house and 58% (105 of 180) sometimes or always drained stagnant water near the house (Fig. 2). Almost all respondents (98%; 177 of 180) would allow their child to be immunized with malaria vaccine, 99% (179 of 180) of respondents are willing to bring their child four times to receive the vaccine and 98% (176/180) would accept malaria vaccination for their child even if given by injection. Most respondents (84%, 151 of 180) would use other preventive measures in addition to malaria vaccination.

**Figure 2.**
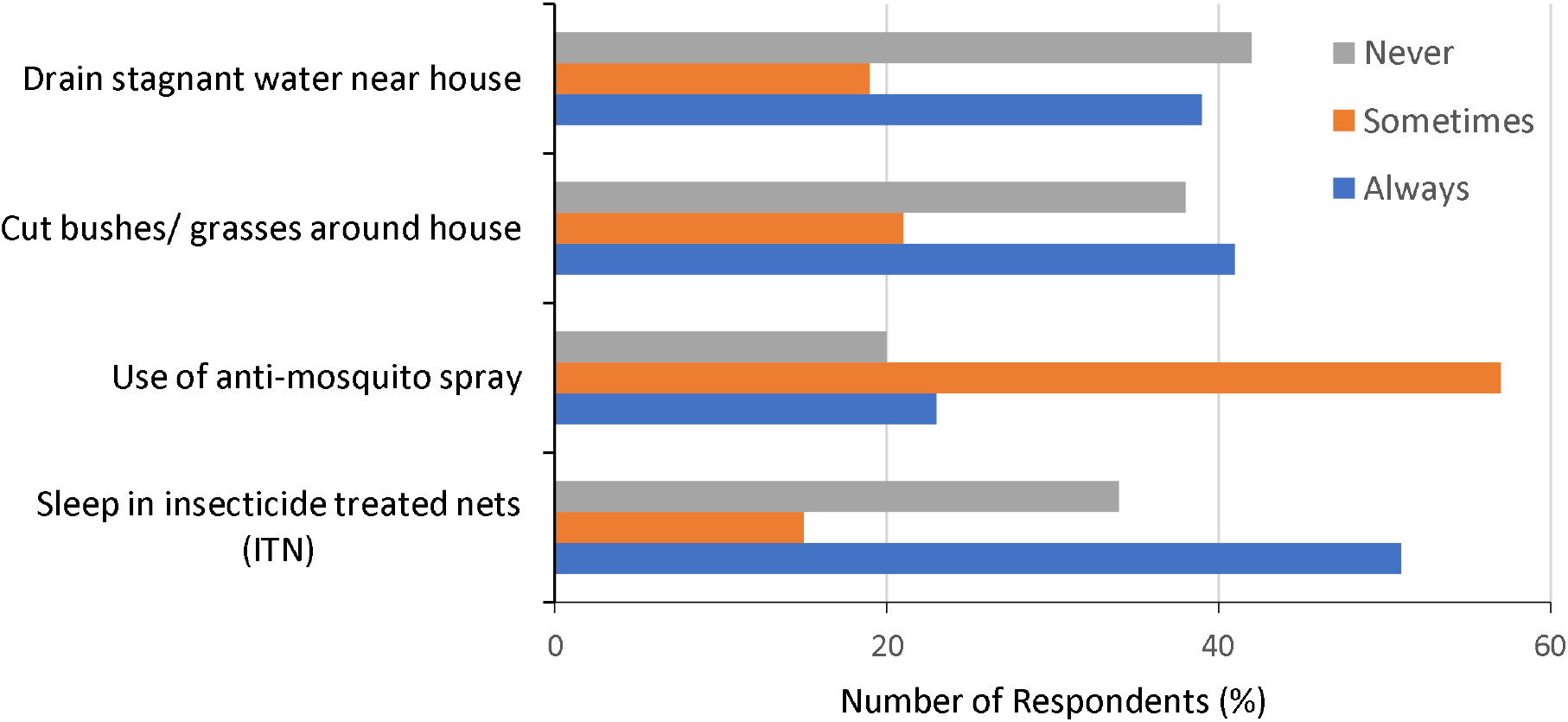
Practice on Malaria Prevention.

## DISCUSSION

With current advances being made on malaria vaccine in the fight against malaria, and the pilot implementation of RTS,S being conducted in three sub-Saharan African countries, a new phase is about to begin in the prevention and control of malaria. To ensure the success of this novel intervention, different factors must be considered, and one is: mothers’ willingness to allow their child(ren) to accept the RTS,S malaria vaccine. Being able to identify mother’s knowledge, attitude, and acceptance of RTS,S is vital in determining steps that need to be taken to ensure optimal uptake and the success of this intervention.

It is evident, from the study, that general knowledge of malaria and its prevention is high. On the other hand, knowledge of malaria vaccine is low and awareness of RTS,S is very low. Factors, identified from this study, influencing knowledge of preventive measures including vaccines are ethnicity, education, age, employment status and number of malaria messages respondents were exposed to. A lot of effort and funds have been invested into behavioral change communication (BCC) materials by various government and non-governmental agencies to increase public awareness of malaria at every level. This could be the reason for the high level of malaria prevention knowledge. This would have to be replicated prior to implementation to attain similar results with RTS,S.

This study found that Hausa and Yoruba mothers were more likely to know three or more preventive measures than Igbos and other tribes. Also, pre-secondary respondents were more likely to know at least 3 preventive measures than post-secondary respondents which means less educated mothers had a better knowledge of malaria prevention. This is not in keeping with previous studies done that suggest that higher education attainment positively influences malaria knowledge.^10,11-14^ The extensive malaria education/training being done at the community level and in health facilities might be a reason for this finding but conducting this research in the other area councils might give a clearer picture.

The study also revealed that receiving three or more malaria messages was found to increase the odds of knowing three or more preventive measures meaning that consistent, repetitive communication of key malaria messages is effective in improving awareness and would be required to increase knowledge of RTS,S when available for routine immunization use. Studies have shown that regular, consistent exposure to BCC messages improve caregivers use of ITNs (a major malaria preventive measure)^15-18^ and is essential for the acceptance of malaria vaccine when available.^10,19^

Furthermore, the study showed that mothers above 30 years were more likely to be aware of malaria vaccines. This could mean that they have had access to more malaria messages, either in the community or health facilities, than the younger respondents. This finding would benefit from further research as a study done in Gwagwalada, showed older age (above 47 years) was associated with poorer knowledge of malaria prevention.^13^

Unemployed respondents were also more likely to know about malaria vaccines than self-employed and formally employed respondents from the results of this study. Although higher wealth index is a factor that determines knowledge of malaria preventive measures,^14^ in this study, being employed did not translate to awareness of malaria vaccine. This finding could suggest that self-employed and formally employed respondents are not exposed to information on public health advances or malaria messages do not include information on malaria vaccine.

Those who had received less than two malaria messaging were more likely to have low attitude scores from this study. This shows that exposure to more malaria messages, irrespective of the source, is necessary to have a positive attitude to malaria prevention. This finding still speaks to the need for constant repetition of malaria messages and novel public health interventions. Although, the Hausas had better knowledge of malaria preventive measures, the knowledge did not translate to a positive attitude towards malaria prevention. This is similar to findings in a study carried out by Singh et al in Aliero, northern Nigeria which showed that knowledge of environmental preventive measures and ownership of ITNs did not necessarily translate to practice of the preventive measures or use of the ITNs.^20^ This was attributed to poor socioeconomic status and low level of formal education within the rural communities.^20^ Igbos with less knowledge of malaria prevention were more likely to have a positive malaria attitude score from this study. With the history of mistrust for western or orthodox medicine among Hausas in Northern Nigeria and the polio vaccination boycott as a precedent,^21^ aggressive advocacy and BCC campaigns will be required to achieve widespread acceptance of RTS,S. This will involve identifying and highlighting positive behaviors to build on, using household, community and political leaders, health facilities and culturally acceptable messages to cultivate the desired attitude and behavior in northern Nigeria.

In addition, the study found that almost all respondents were willing to accept the RTS,S malaria vaccine irrespective of route of administration, efficacy of the vaccine (since they were informed of the efficacy before responding to the practice section) and number of doses their child would have to receive. This is a welcome development showing that the immunization program has done a good job of positively influencing the practices of mothers regarding routine immunization and mothers also understand the severity of the disease. A study by Mbengue et al showed that women who attended antenatal care were more likely to optimally use malaria preventive measures.^22^ Since the women sampled have access to health care facilities and allow their children to be immunized with other routine vaccines, this could account for their willingness to accept the RTS,S malaria vaccine. This might not be the case for mothers in communities who do not have easy access to health care facilities, but this would have to be studied in the future. Also, with the decline in immunizations because of the COVID-19 pandemic,^23^ countries and health communities must identify feasible methods to safely administer vaccines to ensure maximum protection of populations globally.

## CONCLUSION

A lot has been done regarding educating the public on malaria prevention and the benefits of routine immunization. The findings in this study will prove useful to guide policy makers in decision making when RTS,S is approved for routine use. To achieve adequate knowledge, a good attitude and maximum acceptance of RTS,S, messages on the vaccine and its benefits will have to be consistently communicated to the public through various means prior to implementation. Tailoring messages to different audiences based on age, ethnicity, and education will be required. This will ensure optimal uptake of RTS,S and in conjunction with other preventive interventions, bring us one step closer to eliminating malaria.

## Data Availability

No external dataset or supplementary material online at other repositories were used in the manuscript.

## ACKNOWLEDGEMENTS

We acknowledge the respondents who participated in the study. We also acknowledge the contribution of the data collectors, health workers at the public secondary health facilities, and FCT hospital management board. We want to thank the Johns Hopkins Bloomberg School of Public Health (JHSPH) Institutional Review Board (IRB) and the FCT Health Research Ethics Committee for granting approval to conduct this study.

## AUTHORS CONTRIBUTION

TOM conceived the study, designed the study, coordinated data collection and drafted the manuscript, AJA was involved in training of survey personnel and supervision of data collection, BEE analyzed the data and participated in drafting the manuscript, CJS advised on how to conduct the study, draft the manuscript, served as the principal investigator and reviewed the final manuscript. All authors read and approved the final manuscript.

## REFERENCES

1. WHO. Malaria: This year’s World malaria report at a glance. World Health Organization. 2018 November 19 [Cited 2020 July 17]. Available from: https://www.who.int/malaria/media/world-malaria-report-2018/en/

2. WHO. WHO calls for reinvigorated action to fight malaria: Global malaria gains threatened by access gaps, COVID-19 and funding shortfalls. 2020 November 30 [Cited 2020 Decemeber 2]. Available from: https://www.who.int/news/item/30-11-2020-who-calls-for-reinvigorated-action-to-fight-malaria

3. National Malaria Elimination Programme (NMEP), National Population Commission (NPopC), National Bureau of Statistics (NBS), and ICF International. Nigeria Malaria Indicator Survey 2015. Abuja, Nigeria, and Rockville, Maryland, USA: NMEP, NPopC, and ICF International. 2016 [Cited 2020 July 17]. Available from: https://dhsprogram.com/pubs/pdf/MIS20/MIS20.pdf

4. WHO. Malaria: Global Targets. World Health Organization. [Updated 2019 January 25. Cited 2020 July 17]. Available from: https://www.who.int/malaria/areas/global_targets/en/

5. United Nations Sustainable Development Goals Knowledge Platform. Sustainable Development Goal 3. UN. 2016 [Cited 2020 July 17]. Available from: https://sustainabledevelopment.un.org/sdg3

6. World Health Organization. Tables of malaria vaccine projects globally: The Rainbow Tables. WHO. 2015 [Updated 2017 July 17. Cited 2020 July 17]. Available from: http://www.who.int/immunization/research/development/Rainbow_tables/en/

7. WHO. Immunizations, Vaccines and Biologicals: Malaria Vaccines. World Health Organization. 2018 [Cited 2020 July 17]. Available from: https://www.who.int/immunization/research/development/malaria/en/

8. WHO. Malaria: Q&A on the Phase 3 trial results for malaria vaccine RTS,S/AS01. World Health Organization. 2017 April [Cited 2020 July 17]. Available from: https://www.who.int/malaria/media/rtss-phase-3-trial-qa/en/

9. Beliretu I Abdulkadir, Ikeoluwapo O Ajayi. Willingness to accept malaria vaccine among caregivers of under-5 children in Ibadan North Local Government Area, Nigeria. Malaria World Journal 6(2). 2015 [Cited 2020 July 17]. Available from: https://malariaworld.org/mwj/2015/research-willingness-accept-malaria-vaccine-among-caregivers-under-5-children-ibadan-north

10. Chukwuocha UM, Okorie PC, Iwuoha GN, et al. Awareness, perceptions, and intent to comply with the prospective malaria vaccine in parts of South Eastern Nigeria. Malar J. 2018;17(1):187. Published 2018 May 2. doi:10.1186/s12936-018-2335-0

11. Kimbi HK, Nkesa SB, Ndamukong-Nyanga JL, et al. Knowledge and perceptions towards malaria prevention among vulnerable groups in the Buea Health District, Cameroon. BMC Public Health. 2014;14:883. Published 2014 Aug 27. doi:10.1186/1471-2458-14-883

12. Oladimeji KE, Tsoka-Gwegweni JM, Ojewole E, Yunga ST. Knowledge of malaria prevention among pregnant women and non-pregnant mothers of children aged under 5 years in Ibadan, South West Nigeria. Malar J. 2019;18(1):92. Published 2019 Mar 22. doi:10.1186/s12936-019-2706-1

13. Oguntade ES, Shohaimi S, Nallapan M et.al. Factors influencing malaria knowledge, attitude and practice in Gwagwalada. International Journal of Science & Healthcare Research. 2018; 3(3): 168–178.

14. Girum T, Hailemikael G, Wondimu A. Factors affecting prevention and control of malaria among endemic areas of Gurage zone: an implication for malaria elimination in South Ethiopia, 2017. Trop Dis Travel Med Vaccines. 2017;3:17. Published 2017 Dec 20. doi:10.1186/s40794-017-0060-2

15. Zalisk K, Herrera S, Inyang U, et al. Caregiver exposure to malaria social and behaviour change messages can improve bed net use among children in an endemic country: secondary analysis of the 2015 Nigeria Malaria Indicator Survey. Malar J. 2019;18(1):121. Published 2019 Apr 6. doi:10.1186/s12936-019-2750-x

16. Bowen HL. Impact of a mass media campaign on bed net use in Cameroon. Malar J. 2013;12:36. doi: 10.1186/1475-2875-12-36.

17. Owusu Adjah ES, Panayiotou AG. Impact of malaria related messages on insecticide-treated net (ITN) use for malaria prevention in Ghana. Malar J. 2014;13:123. doi: 10.1186/1475-2875-13-123.

18. Elmosaad YM, Elhadi M, Khan A, et al. Communication for behavioural impact in enhancing utilization of insecticide-treated bed nets among mothers of under-five children in rural North Sudan: an experimental study. Malar J. 2016;15:509. doi: 10.1186/s12936-016-1551-8.

19. Ojakaa, D.I., Ofware, P., Machira, Y.W. et al. Community perceptions of malaria and vaccines in the South Coast and Busia regions of Kenya. Malar J 10, 147 (2011). https://doi.org/10.1186/1475-2875-10-147

20. Singh R, Musa J, Singh S, Ebere UV. Knowledge, attitude and practices on malaria among the rural communities in aliero, northern Nigeria. J Family Med Prim Care. 2014;3(1):39–44. doi:10.4103/2249-4863.130271

21. Jegede AS. What led to the Nigerian boycott of the polio vaccination campaign?. PLoS Med. 2007;4(3):e73. doi:10.1371/journal.pmed.0040073

22. Mbengue MAS, Bei AK, Mboup A, et al. Factors influencing the use of malaria prevention strategies by women in Senegal: a cross-sectional study. Malar J. 2017;16(1):470. Published 2017 Nov 21. doi:10.1186/s12936-017-2095-2

23. WHO. WHO and UNICEF warn of a decline in vaccinations during COVID-19. World Health Organization. 2020 July 15 [Cited 2020 August 4]. Available from https://www.who.int/news-room/detail/15-07-2020-who-and-unicef-warn-of-a-decline-in-vaccinations-during-covid-19

